# Real-world federated learning for the brain imaging scientist

**DOI:** 10.1101/2023.04.22.23288741

**Authors:** Stijn Denissen, Jorne Laton, Matthias Grothe, Manuela Vaneckova, Tomáš Uher, Matěj Kudrna, Dana Horáková, Johan Baijot, Iris-Katharina Penner, Michael Kirsch, Jiří Motýl, Maarten De Vos, Oliver Y. Chén, Jeroen Van Schependom, Diana Maria Sima, Guy Nagels

**Author notes:** Correspondence: Stijn Denissen (, Twitter: @StijnDenissen).

## Abstract

**Background:** Federated learning (FL) could boost deep learning in neuroimaging but is rarely deployed in a real-world scenario, where its true potential lies. Here, we propose FLightcase, a new FL toolbox tailored for brain research. We tested FLightcase on a real-world FL network to predict the cognitive status of patients with multiple sclerosis (MS) from brain magnetic resonance imaging (MRI).

**Methods:** We first trained a DenseNet neural network to predict age from T1-weighted brain MRI on three open-source datasets, IXI (586 images), SALD (491 images) and CamCAN (653 images). These were distributed across the three centres in our FL network, Brussels (BE), Greifswald (DE) and Prague (CZ). We benchmarked this federated model with a centralised version. The best-performing brain age model was then fine-tuned to predict performance on the Symbol Digit Modalities Test (SDMT) of patients with MS (Brussels: 96 images, Greifswald: 756 images, Prague: 2424 images). Shallow transfer learning (TL) was compared with deep transfer learning, updating weights in the last layer or the entire network respectively.

**Results:** Centralised training outperformed federated training, predicting age with a mean absolute error (MAE) of 6.00 versus 9.02. Federated training yielded a Pearson correlation (all p < .001) between true and predicted age of .78 (IXI, Brussels), .78 (SALD, Greifswald) and .86 (CamCAN, Prague). Fine-tuning of the centralised model to SDMT was most successful with a deep TL paradigm (MAE = 9.12) compared to shallow TL (MAE = 14.08), and respectively on Brussels, Greifswald and Prague predicted SDMT with an MAE of 11.50, 9.64 and 8.86, and a Pearson correlation between true and predicted SDMT of .10 (p = .668), .42 (p < .001) and .51 (p < .001).

**Conclusion:** Real-world federated learning using FLightcase is feasible for neuroimaging research in MS, enabling access to a large MS imaging database without sharing this data. The federated SDMT-decoding model is promising and could be improved in the future by adopting FL algorithms that address the non-IID data issue and consider other imaging modalities. We hope our detailed real-world experiments and open-source distribution of FLightcase will prompt researchers to move beyond simulated FL environments.

## Introduction

Deep learning is gaining traction as a tool to study the brain’s function and structure (1,2). Since the comprehensive Nature review by Yann Lecun, Yoshua Bengio and Geoffrey Hinton on the topic in 2015 (3), the curiosity of brain researchers has clearly been sparked; over 10.000 papers were published on the topic in the last decennium, while just over 600 were published at the end of 2014.

Deep learning has led to major breakthroughs in brain analysis. Brain structures can now be accurately quantified using segmentation models (4,5), aiding doctors in diagnosis and evaluating treatment effect. Deep neural networks furthermore reduce the error in predicting age from brain MRI to the order of 2 years (6). This yields accurate biological aging clocks that capture deviations from healthy aging patterns and efficient communication tools; brain damage can be expressed in terms of “how much older the brain looks”. Deep learning models can indeed capture the brain’s complexity by creating high-dimensional, data-driven representations beyond human understanding. Yet there is a catch. To create reliable representations, deep learning models require big training datasets; usually in the order of tens of thousands of images (6). As many dedicated researchers put significant effort in collecting datasets, we need to reconsider how they can be reused and combined to unlock the full potential of deep learning in brain image analysis.

The conventional way to train deep learning models is to centralize datasets. This is feasible for certain domains like brain age, as it relies on healthy control data sets that are publicly shared. Indeed, age-labelled T1-weighted images are widely available via initiatives such as the UK Biobank (7) and many repositories in OpenNeuro (8). In most other domains however, especially when working with sensitive patient data, data sharing is difficult. Barriers to data sharing can be plentiful in technical, motivational, economic, political, legal, and ethical terms (9). Especially barriers in the latter three domains apply for centralizing neuroimaging data. A first conditio sine qua non is trust; “In the absence of trust, providers could anticipate potential misinterpretation, misuse or intentional abuse of the data” (9). Second, the Global Data Protection Regulation (GDPR) enforces strict guidelines to data sharing. This either complicates the procedure, lowering the incentive to do so, or blocks it entirely. This is especially true for the type of data brain scientists work with, as faces could be reconstructed from MR images. Lastly, data sharing is relatively static. Especially data from routine clinical practice is continuously generated, and sharing those periodically is time-consuming and inefficient.

This conventional, centralized view on machine learning was challenged by McMahan and colleagues in 2017 (10). They introduced the concept of Federated Learning (FL), in which a model is trained by sending it to the data in local institutions, where they are trained locally. Instead of data, models are shared. Moreover, the computational load and data storage is spread across multiple centres. In the following years, (brain) researchers started experimenting with the idea in simulated settings, mainly on open-source data. New algorithms were suggested (11), performance with respect to centralised training was explored (12) and federated learning toolboxes were designed (13). Yet while FL in principle solves all previously mentioned issues, it generates new challenges that hinder brain scientists to deploy it in a real-world setting. Among those are financial boundaries (e.g. graphical processing units (GPU)), hardware and software differences, connectivity issues and data heterogeneity. Indeed, only few succeeded to date, mainly in brain tumour segmentation (14,15), where access to bigger datasets boosted model performance (15). While other real-world examples exist, it is often unclear from the methods section of papers whether it concerned a simulation or real-world FL on geographically distributed data.

In this work, we aspired to pioneer real-world FL in our own modelling domain, decoding cognitive performance from structural brain MRI in multiple sclerosis (MS) on data from three international centres in Brussels (BE), Greifswald (DE) and Prague (CZ). When starting the practical setup in early 2023, the most pressing issue was the lack of software orchestrating real-world federated learning in our neuro-imaging context, not to mention handling data loading of clinical datasets that differed widely in format. Hence, we designed a simple, glass-box FL framework that allows easy real-world deployment in an international context: “FLightcase”. FLightcase is designed to work with the Brain Imaging Data Structure (BIDS (16)), which became the standard data organisation format for brain researchers in the past decade (17). BIDS is enforced by data sharing platforms such as OpenNeuro (8), and we aspired to continue this trend. In this way, we hope to encourage brain AI researchers to consider decentralised model training and leverage their modelling to larger, multi-institutional datasets to boost generalisability.

The contributions of this paper are therefore the following:

- The introduction of FLightcase, a simple, open-source, BIDS-compliant FL toolbox in neuroimaging.
- Proving the real-world readiness of FLightcase on a real-world FL network with geographically distributed data in Brussels (BE), Greifswald (DE) and Prague (CZ). The modelling goal was to predict cognitive function from brain MRI in MS using transfer learning from a pre-trained brain age model. In doing so, we addressed two research questions:
  1. Does federated match centralised training in terms of model performance? This question was addressed in the brain age modelling, as it relies on open-source data that can be centralised.
  2. Does deep transfer learning (updating all network weights) outperform shallow learning (updating only the weights of the fully connected layer) for predicting cognitive impairment in MS?
- Discussing challenges and solutions in setting up a real-world federated learning network to encourage the method in the field.

## Methods

### FLightcase in brief

FLightcase is a federated learning toolbox that was specifically designed to work with the Brain Imaging Data Structure (BIDS) (16). We thereby aim to facilitate model training for brain researchers and stimulate researchers to use this data structure. Its basic communication relies on sending 2 files after each other. The first contains the machine learning information to be transmitted, the second is a text file marking transmission completion. Files are sent between computers via secure copy protocol (SCP) and therefore require all computers to be UNIX-based. In the remainder of this paper, we will refer to “computer” or “node” interchangeably. The SCP command requires two localisers:

- The IP address of the receiving computer. An example of a secure network where computers are assigned an IP address, and are therefore reachable, is a virtual private network (VPN). This was used in our real-world example (cfr. infra).
- The location within the computer to write the file to. To facilitate this, we use the concept of an “FL workspace”. The FL workspace is a folder dedicated to the federated learning experiment.

These and other settings (cfr. infra) are stored in a JavaScript Object Notation (JSON) file per computer, which are different for clients and server. For example, the client settings define the location of the BIDS dataset, the server settings define the clients to expect. To orchestrate the FL process, the server moreover stores an “FL plan” JSON file containing training preferences (e.g. number of FL rounds). Lastly, the server defines the model architecture that will be trained in a separate Python file.

For a more in-depth overview of the FLightcase software, we refer to the “FLightcase unpacked” section of the supplementary material. FLightcase is publicly available in our AIMS-VUB GitHub repository (https://github.com/AIMS-VUB/FLightcase) and deployed on the Python Package Index (PyPI, https://pypi.org) for easy installation. The federated experiments in this study relied on FLightcase v0.1.8. FLightcase can be tested using the simulation explained in the supplementary material, section “How to test FLightcase”.

FLightcase v0.1.8 depends on the following Python modules: torch v2.5.1 (18), pandas v2.2.3 (19), monai v1.4.0 (20), scikit-learn v1.6.0 (21), tqdm v4.67.1 (22), nibabel v5.3.2 (23), paramiko v3.5.0 (24), scp v0.15.0 (25), matplotlib v3.10.0 (26), scipy v1.14.0 (27), numpy v1.26.4 (28), click v8.1.8 (29), twine v6.0.1 (30).

### The federated learning network

To prove the real-world effectiveness of FLightcase, we tested it in an FL network between our AIMS lab in Brussels (BE), the Greifswald University Hospital (DE) and the General University Hospital Prague (CZ). The network (figure 1) consists of 4 computers, of which one is the server that coordinates the project, and the other three serve as clients on which models are trained using the local data. The two Brussels computers were in the same office and connected to the network of the department of electronics and informatics (ETRO) of the VUB. The computers in Greifswald and Prague were connected to this network via a Virtual Private Network (VPN). Models were shared via secure copy protocol (SCP) with secure shell (SSH).

**Figure 1.**
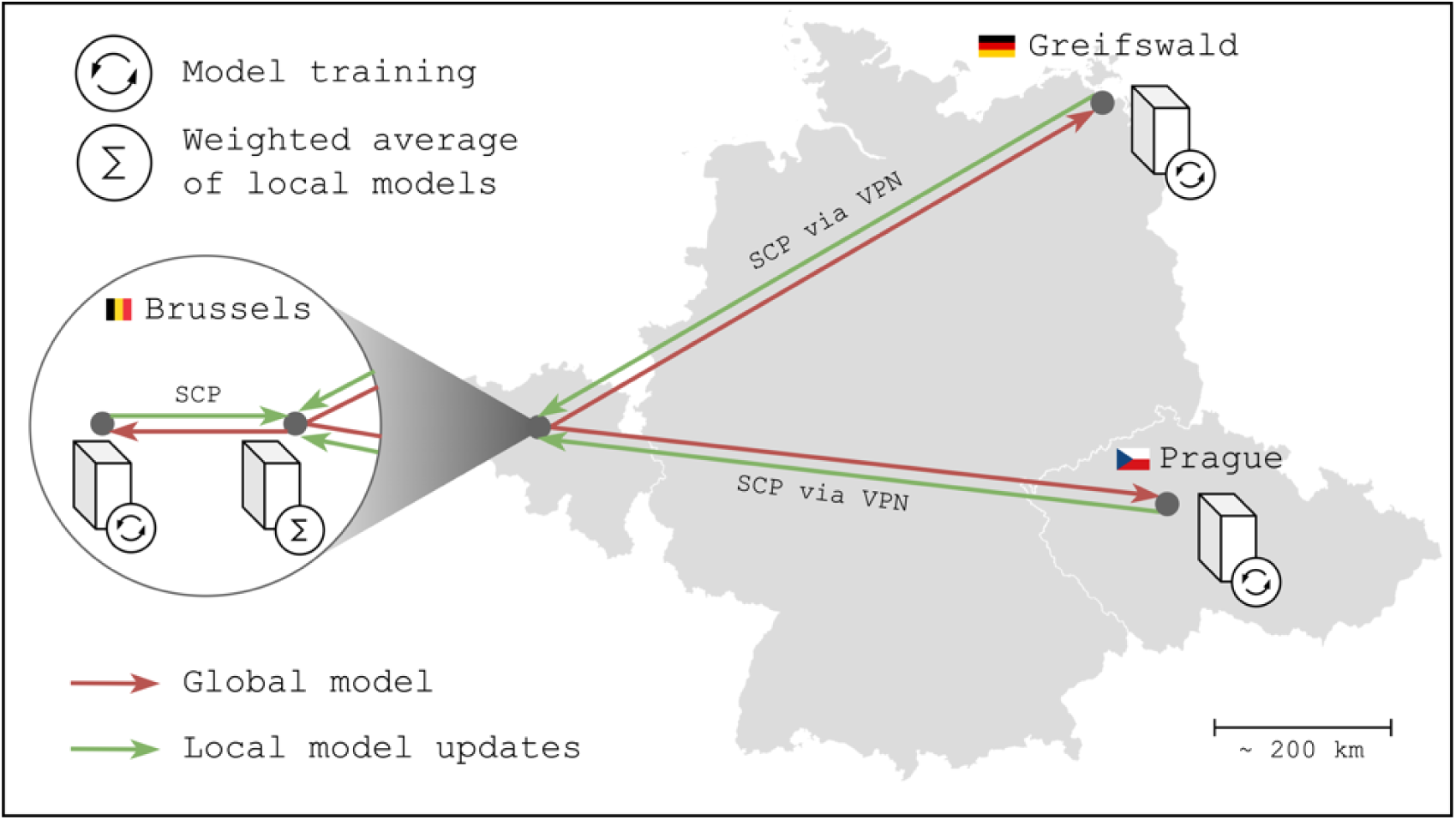
The Federated Learning network for the real-world experiment. The computer with the “sigma” symbol is the server, whereas computers with an “update” symbol are clients. Abbreviations: SCP = Secure Copy Protocol; VPN = Virtual Private Network.

All client computers were equipped with a graphical processing unit (GPU); Brussels: NVIDIA GeForce RTX 4090 (24GB), Greifswald: NVIDIA GeForce RTX 3090 (24GB) and Prague: NVIDIA GeForce RTX 4090 (24GB). The operating system of each computer (server and clients) was Debian GNU/Linux 12 (“bookworm”), and Python version 3.11.2 was used consistently.

### Real-world FLightcase testing

This section explains the two steps to test FLightcase on the real-world FL network explained above. Step 1 involved training a DenseNet convolutional network architecture (31) to predict age from T1-weighted brain MRI, analogous to the work of James Wood and colleagues (32). As step 1 modelled on open-source data and thus allowed centralising data, federated training was benchmarked against centralised training (explained at the end of the methods section). Step 2 involved transfer learning of the best brain age model from step 1 (lowest overall test mean absolute error (MAE)) to predict cognitive impairment in people with multiple sclerosis (MS). The FL plans for training the brain age and SDMT models were included as supplementary table S4.

### Ethics

The “Commissie Medische Ethiek” (CME) of the UZ Brussel judged this retrospective study to be exempt from ethical approval (B.U.N. 1432022000303). For the MS data at each center in this study, ethical approval was obtained prior to data acquisition (Brussels: B.U.N. 143201423263, Greifswald: BB159/18, Prague: 113/22 S-IV and 28/17). Healthy control data used to train the brain age model was obtained from open-source datasets.

### Data

To train the brain age network (step 1), we used three open-source data sets: the Cambridge Centre for Ageing and Neuroscience (Cam-CAN) dataset (33), the Information eXtraction from Images (IXI) dataset (34) and the Southwest University Adult Lifespan Dataset (SALD) (35). The Cam-CAN dataset was stored on the Prague computer, the IXI dataset in Brussels and the SALD dataset in Greifswald. The datasets contain T1-weighted MRI, sex and age from healthy subjects. The data is summarised in table 2.

**Table 2.**
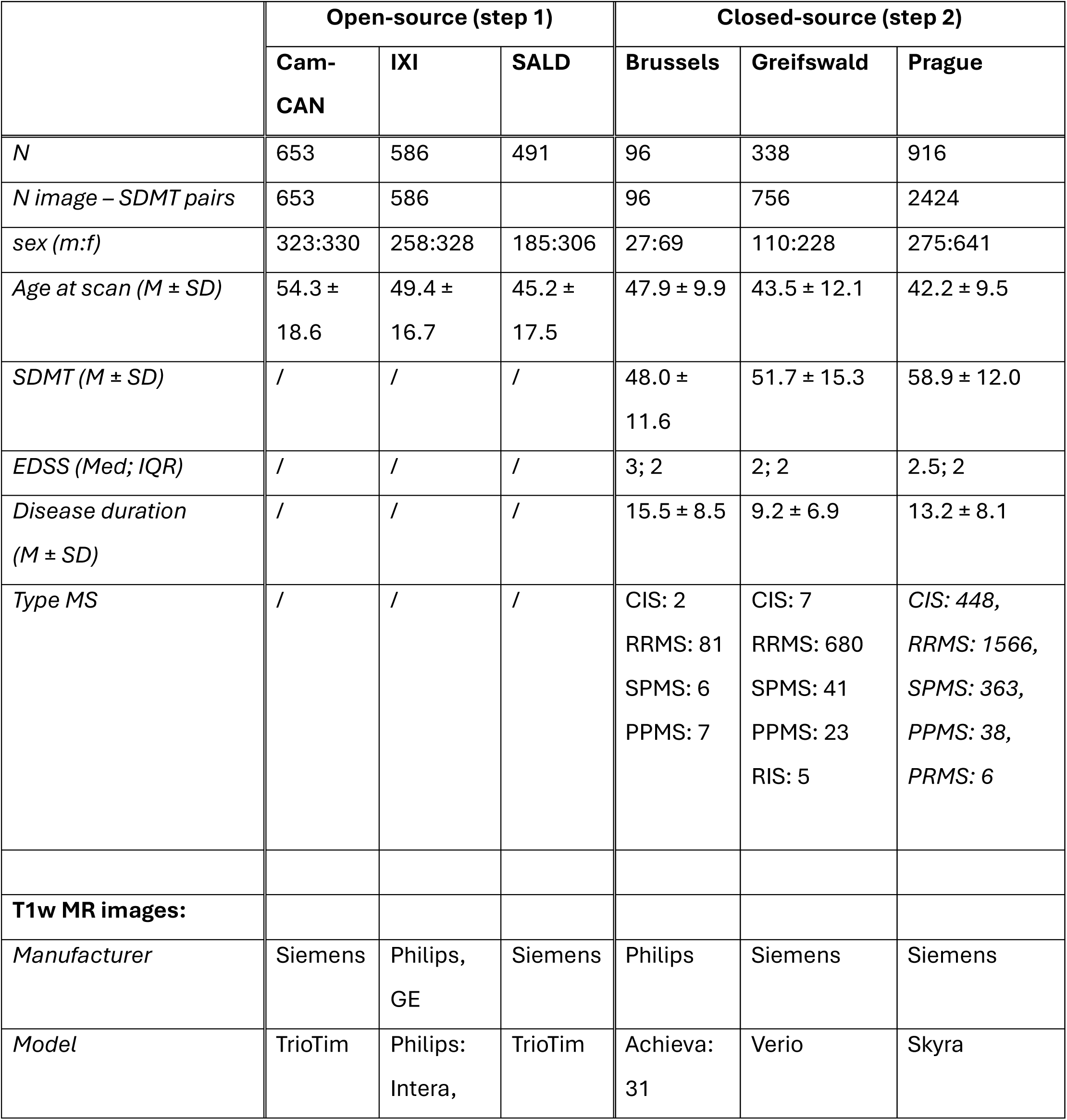
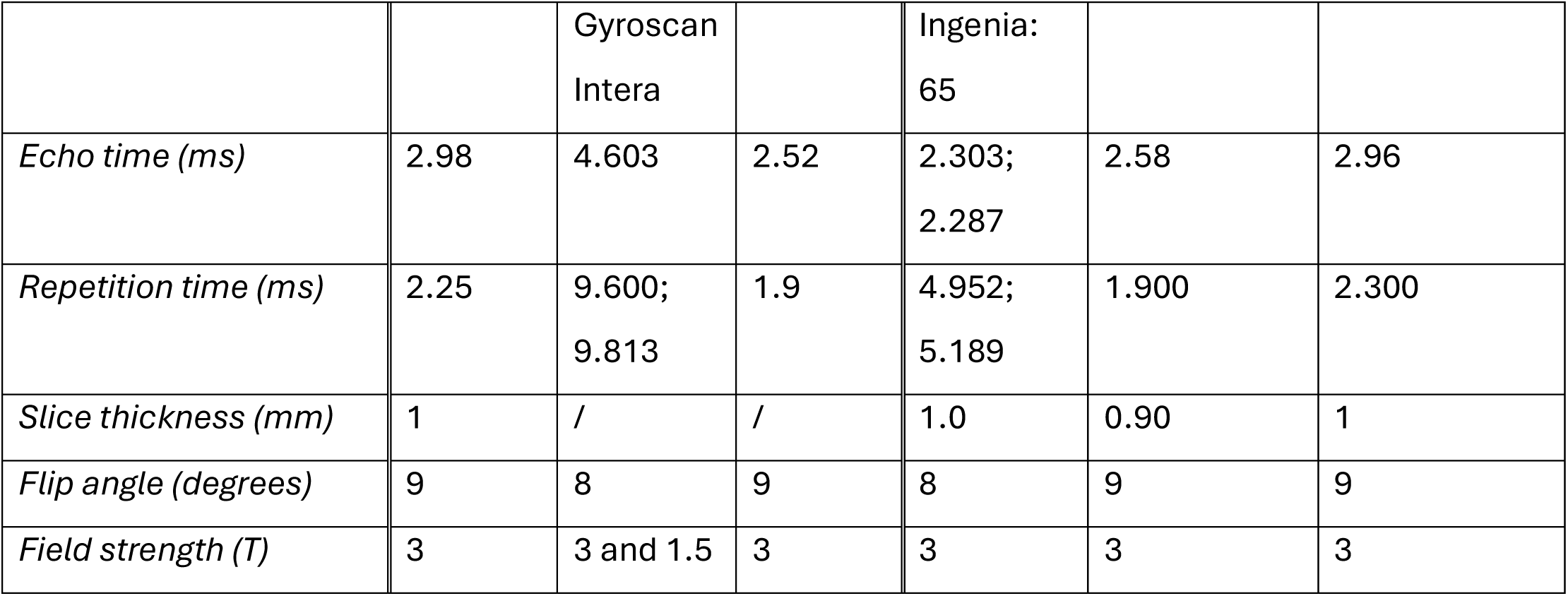
Dataset characteristics. Abbreviations: n = sample size, m = male, f = female, M = mean, SD = standard deviation, SDMT = symbol digit modalities test, EDSS = expanded disability status scale, GE = General Electric. Notes: [1] Variable distributions are calculated across all image-SDMT pairs. [2] missing values: 14 EDSS (Prague), 3 disease course (Prague) and 15 disease duration (Greifswald). [3] The MR image acquisition info was extracted from a single subject per scanner model and might therefore deviate among subjects. For IXI, which was acquired at three different sites, M RI acquisition info was obtained from the website: https://brain-development.org/ixi-dataset/ (accessed 10 March 2025). Detailed scanner info was only available for the Philips scanners.

The cognition prediction network (step 2) was trained on three MS datasets at the Vrije Universiteit Brussel (VUB), the general university hospital Prague (VFN) and Greifswald university hospital. The data were organised locally in the BIDS format, and contained T1 weighted MR images, demographic and clinical information. This entailed sex, age, expanded disability status scale (EDSS (36), physical disability), disease duration, MS subtype (relapsing versus progressive onset) and the symbol digit modalities test (SDMT (37)), i.e., the target to predict. In this test, a subject is presented a list of symbols that need to be converted to numbers using a key on the top of the page, matching symbols with numbers. In 90 seconds, the subject must convert as many symbols to numbers as possible, each time saying the number out loud for the test administrator to write down. The SDMT is a measure of information processing speed.

The T1-weighted MR images were preprocessed using the pipeline by Wood et al. 2022 (32), which includes skull-stripping, registration to Montreal Neurosciences Institute (MNI) 152 space (1mm isotropic) and cropping to an isotropic resolution of 130 voxels. These steps are defined in their “pre_process.py” script, for which we included an updated version in our GitHub repository with permission of the authors (32).

The data was split randomly in 80% train and 20% test data on each client. To prevent data leakage, multiple images of a single subject were collected in either train or test set. Each FL round, the training data was moreover bootstrapped 5 times, with 75% train and 25% validation data. This results in a train/validation/test split of 60/20/20 (cfr. supplementary table S4).

### The 3D DenseNet model

The 3D Dense Convolutional Network (DenseNet) (31) was used during the initial brain age task (step 1) and the transfer learning task to SDMT (step 2). DenseNet outperformed other network architectures in a medical imaging context (38) and is unique for directly connecting all layers inside the network with each other (31). Each layer therefore takes all previous feature maps, outputs of previous layers, as input. Hence, the propagation of features throughout the network is improved. The DenseNet architecture moreover reduces the “vanishing gradient” problem, where the gradient used to update weights in the network gradually approaches zero during backpropagation to earlier layers. Lastly, it reduces the number of parameters in the network (31). The 3D DenseNet model in this paper has 11243649 updatable parameters and is described schematically in figure 2.

**Figure 2.**
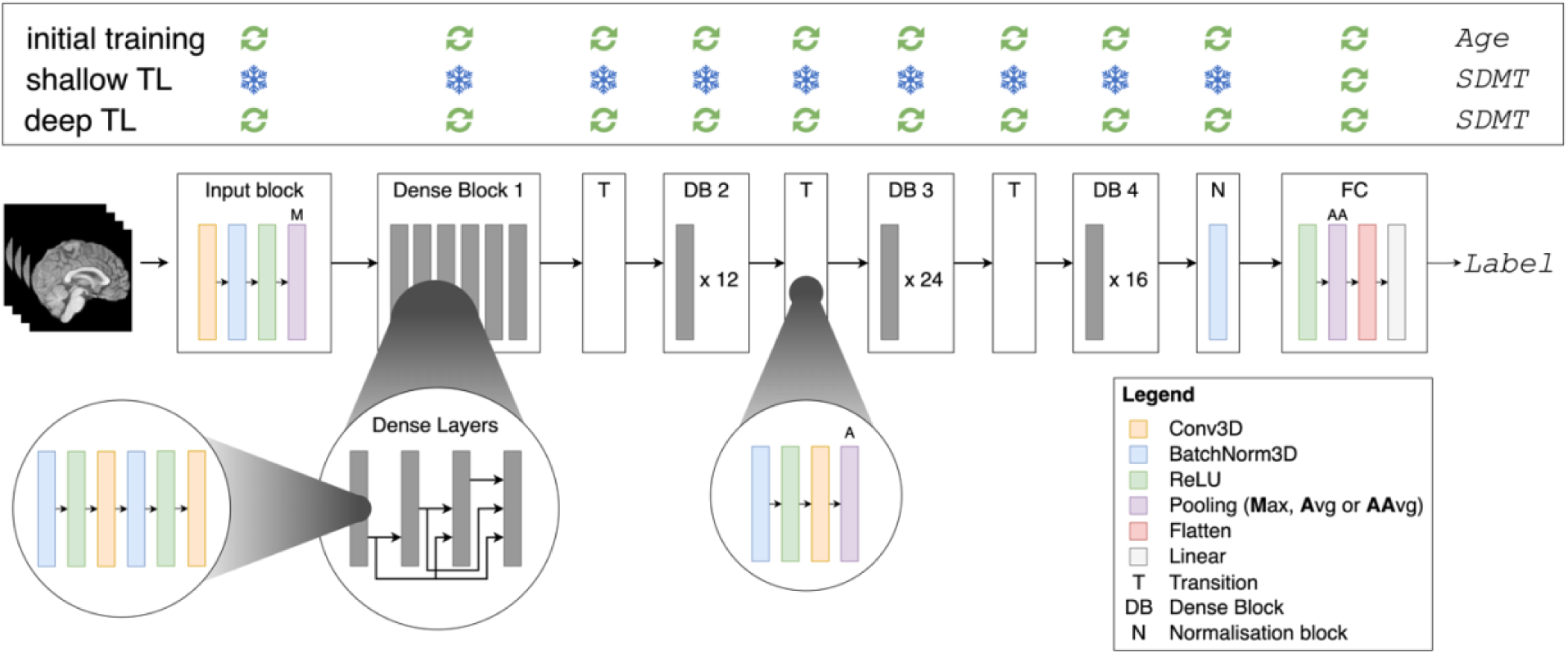
DenseNet architecture and training methodology (top panel). Avg = Average, AAvg = Adaptive Average. The “Label” is chronological age for the initial brain age prediction task, and the SDMT for the shallow and deep transfer learning (TL).

For the brain age prediction task, all layers in the network were unfrozen, whereas for the SDMT prediction task, two transfer learning methods were explored. In the “shallow TL” task, only the fully connected layer, consisting of 1025 parameters (1024 weights and 1 bias), was updated during training (cfr. figure 2); the other layers, i.e. the feature extractor part of the network, was frozen. In the “deep TL” task, all layers of the network were updated. Figure 2 also summarises the transfer learning methodology in the box on top.

### Evaluation

The final performance of all models was evaluated on the test data set per client by the mean absolute error (MAE) and Pearson correlation. The overall model performance was moreover calculated with equation 1:

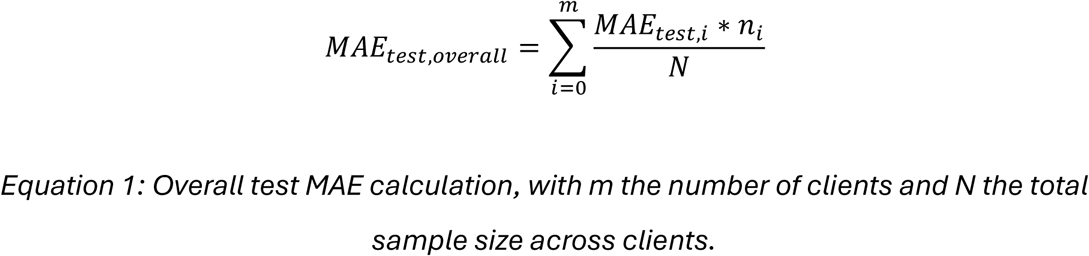

### Benchmarking: centralised brain age training

As a benchmark model, we additionally updated the brain age model on a centralised version of the three open-source datasets. The same test dataset was used as in the federated setting. During training however, the random train/validation splits (5 per epoch) were performed on the entire training dataset (80%). The training dataset therefore included samples of all three datasets, while these were separated in the federated setting. As the centralised setting had access to all test results together, the test MAE was calculated across all subjects simultaneously instead of the per-client approach in equation 1.

## Results

### Step 1: predicting brain age

#### Real-world federated training

The total FL process took 2 hours, 22 minutes, and 50 seconds to complete. The model reached a minimum after 48 rounds and training stopped early after 68 rounds. The final network achieved an overall test MAE of 9.02, and an MAE of 8.86, 8.45 and 9.92 on the test datasets of CamCAN, IXI and SALD respectively. Pearson correlation for the respective datasets was 0.78 (p < .001), 0.78 (p < .001) and 0.86 (p < .001). The training process is visualised in figure 3, whereas scatterplots between true and predicted age (brain age) are displayed in figure 4.

**Figure 3:**
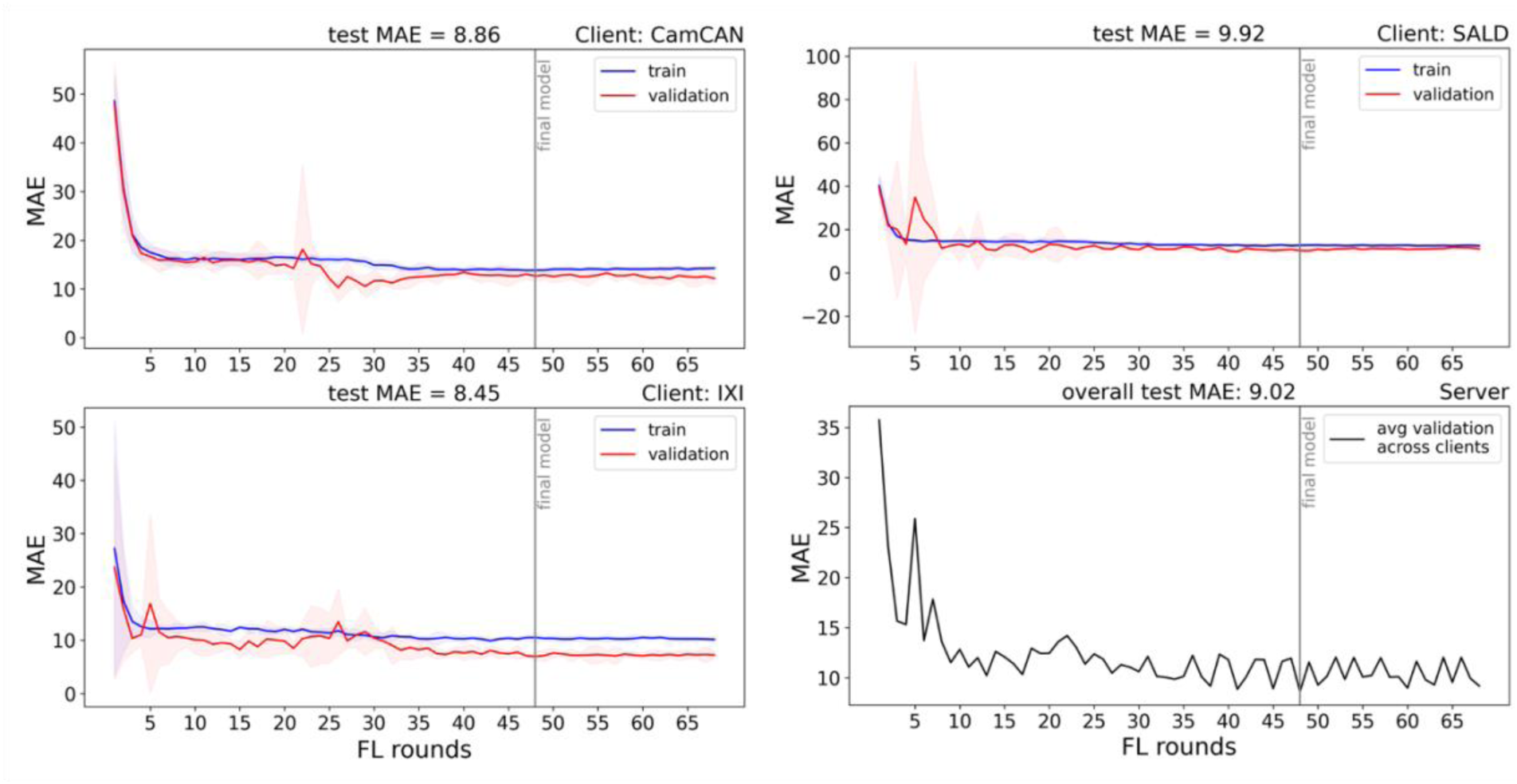
Federated training of the brain age network. The red (train) and blue (validation) lines represent the average train and validation MAE respectively, and the shaded red and blue areas represent the 95% confidence interval, all calculated across 5 bootstraps per FL round. The right lower panel show the average validation MAE across the clients in the aggregation, calculated on the server side.

**Figure 4.**
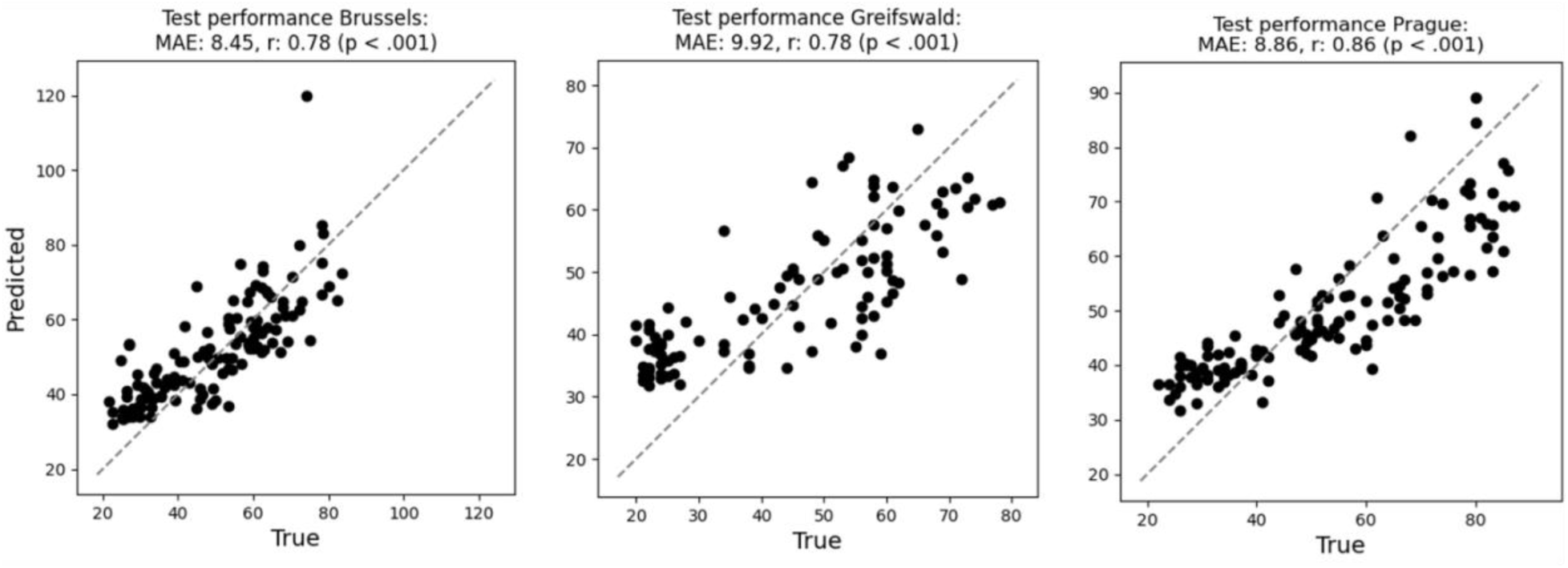
Scatterplots between true and predicted age (brain age) on the test datasets of CamCAN (stored in Prague), IXI (stored in Brussels) and SALD (stored in Greifswald). The grey dashed line indicates the perfect prediction line where Age = Brain Age.

#### Centralised benchmark

Centralised training on the three open-source data sets took 10 hours and 8 seconds. The model reached a minimum after 108 rounds and was stopped early after 128 rounds. The model achieved an overall test MAE of 6.00 and a Pearson correlation of 0.91 (p < .001). The results are displayed visually in supplementary figure S2 (loss figure) and S3 (test results scatterplot).

### Step 2: predicting SDMT

Figure 5 shows the training process for training the SDMT models using shallow TL (left) and deep TL (right), starting from the centralised brain age model. Figure 6 shows the results of applying the final model to the test set of each centre.

**Figure 5.**
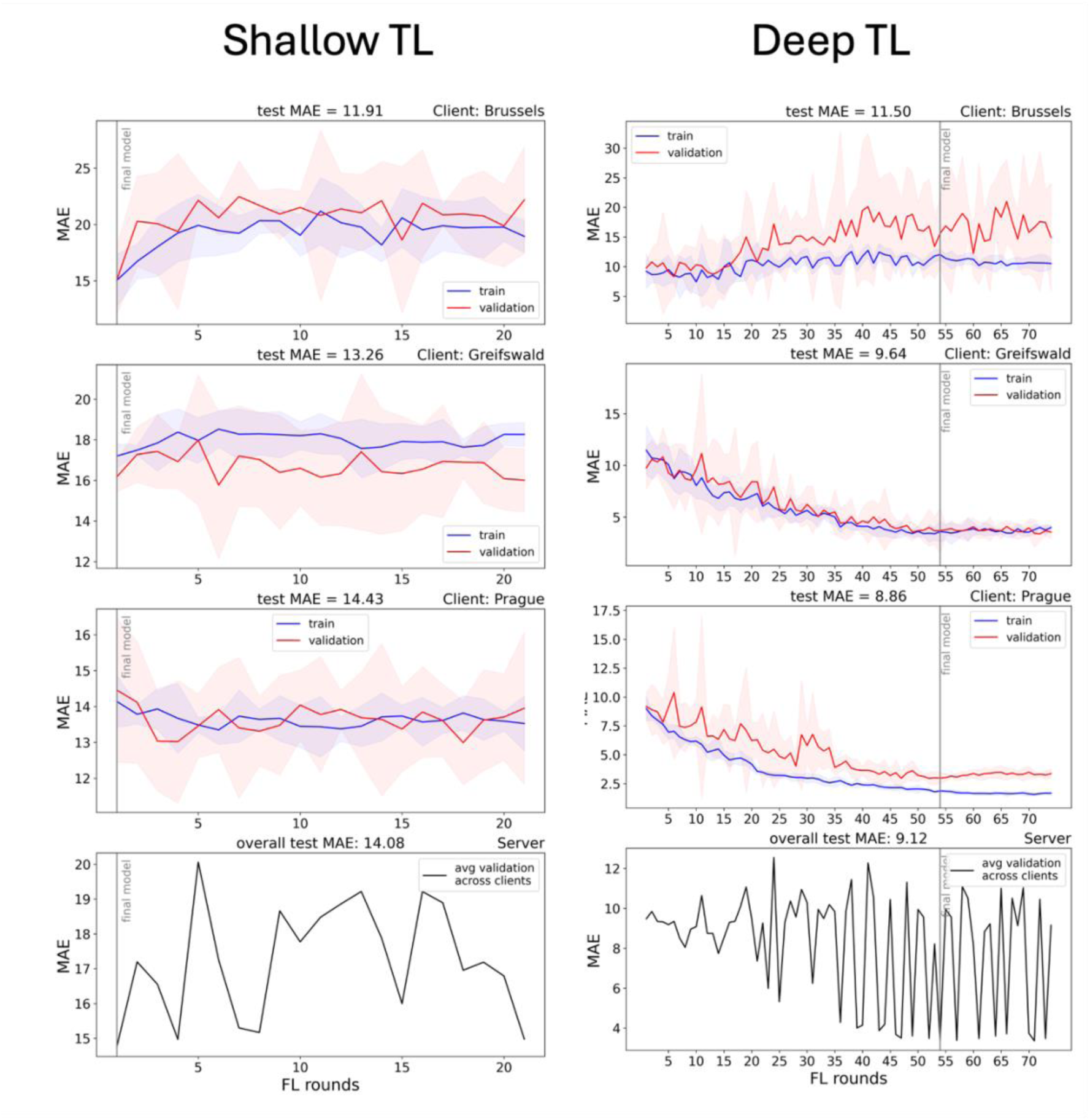
Loss figure of fine-tuning the brain age model to predict SDMT using shallow (left) and deep (right) transfer learning. Abbreviations: FL = federated learning, MAE = mean absolute error, avg = average. The lower panels show the average validation MAE across the clients in the aggregation, calculated on the server side.

**Figure 6.**
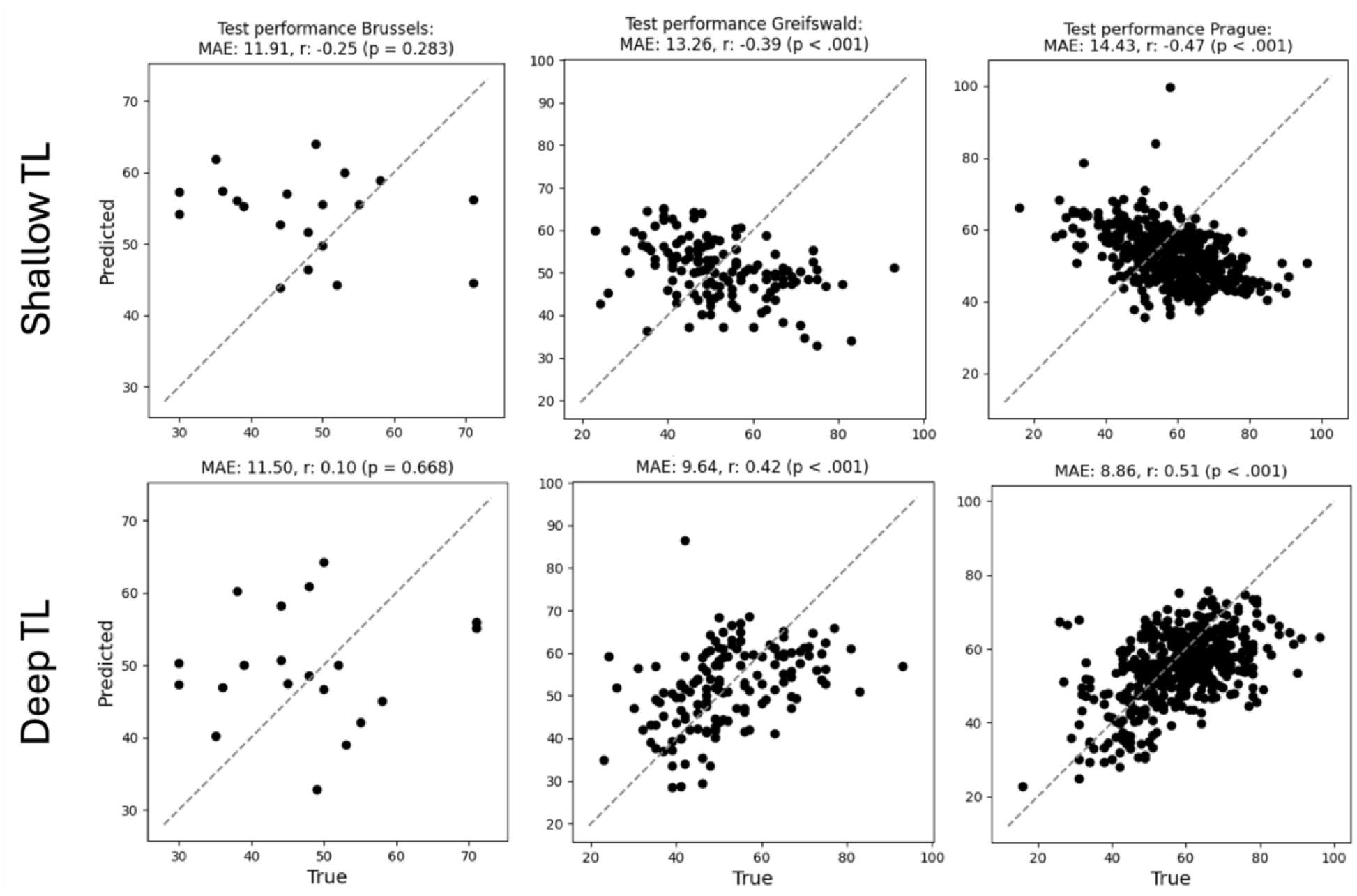
Scatterplots visualising test performance of the SDMT model trained with shallow TL (upper row) and deep TL (lower row).

Shallow TL took 1 hour, 49 minutes and 42 seconds. The model reached a minimum average validation MAE after the first FL round and achieved an overall test MAE of 14.08 SDMT points. The per-client test performance in terms of MAE was 11.91 (Brussels), 13.26 (Greifswald) and 14.43 (Prague), whereas Pearson correlation was –0.25 (p=0.283), –0.39 (p<.001) and –0.47 (p < .001) respectively.

For the deep TL model (right), training took 8 hours, 30 minutes and 4 seconds. The model reached a minimum at FL round 54 and achieved an overall test MAE of 9.12 SDMT points. The per-client test performance in terms of MAE was 11.50 (Brussels), 9.64 (Greifswald) and 8.86 (Prague), whereas Pearson correlation was 0.10 (p=0.668), 0.42 (p<.001) and 0.51 (p < .001) respectively.

## Discussion

In this manuscript, we introduced FLightcase, a Federated Learning toolbox specifically designed to promote real-world, decentralised machine learning for brain scientists. We then proved the real-world readiness of FLightcase by training models to predict SDMT from T1-weighted brain MRI images, using transfer learning from a brain age model. During brain age modelling, centralised training outperformed federated training. During transfer learning, a deep paradigm where all network weights were trained outperformed shallow learning, where solely the last network layer was updated. The final model predicted SDMT with an average error of 9.12 points and performed especially well on the Greifswald and Prague dataset.

### Decentralised brain age modelling

The final federated brain age model predicted age from T1w MRI of healthy controls with an average error of 9.02 years across datasets. Strong correlations were furthermore observed on each client test dataset, indicating that federated learning successfully sensitised the model to individual differences in brain structure. Nonetheless, centralised training outperformed the federated approach with a test MAE reduction of 3.02 years. This contrasts findings from an FL simulation by Basodi et al. 2022, finding similar performance of brain age models trained on decentralised and centralised datasets (12). Besides a larger dataset of over 10.000 images from 2 sources, data from each source was distributed across 6 simulated centres, contrasting our design with one source per centre.

Our brain age models might suffer low generalisability by nature of the open-source datasets, containing both Caucasian (Cam-CAN and IXI) and Chinese (SALD) brains that are known to differ in structure (39,40). The model indeed had a higher test MAE in SALD (9.92) compared to CamCAN (8.86) and IXI (8.45). The overfitting behaviour on the IXI dataset was moreover visible in the average validation loss in the aggregation sample at the server side; valleys coincided with IXI being in the sample. This zigzagging pattern was especially clear in the last rounds (figure 3). This underlines the importance to consider alternative client sampling schemes such as FedSampling (41). Lastly, the combined size of the decentralised dataset was 1730 images, while the best model reported in a comprehensive review on brain age model performance (6) was trained by Han Peng and colleagues on 14.503 T1-weighted brain images (42). Enlarging our dataset would likely have resulted in improved brain age prediction performance, as well as a lower discrepancy between centralised and federated training such as in Basodi et al. 2022 (12).

### Transfer learning to SDMT

Deep transfer learning from the centralised brain age model outperformed shallow transfer learning with an overall MAE of 9.12 versus 14.08. This indicates that the latent feature representation for the brain age task was incompatible with the SDMT prediction task, requiring an update of the feature extractor part of the network.

Although deep TL indeed improved performance substantially, the model had difficulty decoding SDMT performance on the Brussels dataset. While training and validation loss as well as confidence intervals gradually decreased in Greifswald and Prague, loss increased in the Brussels dataset. Not unlike federated brain age modelling (cfr. supra), this translates in heavy loss fluctuations on the server side, depending on which centre models are in the aggregation sample. Valleys coincided with the local models of Greifswald and Prague in the aggregation sample. We hypothesise two factors to underly this behaviour.

First, the Greifswald and Prague datasets were larger. The contributions of their local models in each round were therefore bigger than the Brussels local model, causing the model to learn less from the Brussels dataset. This is an inherent problem of the conventional FedAvg paradigm, which is why solutions for equality of local model updates start to emerge. An example of this is q-FedAvg, which “reweights” the loss by a parameter q (43). The result is that client models with higher loss receive a higher weight in the aggregation.

Second, the behaviour nicely illustrates the non-IID problem in federated learning, meaning that we cannot assume that data points are drawn from the same underlying sample (44). This violates an important assumption in machine learning (45). Indeed, MS centres work with different equipment and workflows, which can be expected to increase across country borders. As illustrated in table 1, different MRI scanners were used across different centres, and MS samples differed, most prominently in the SDMT ground truth distribution. This raises awareness for the importance of data harmonisation and gaining insights in between-centre differences in for example test administration. It is therefore encouraging to observe the emergence of open-source federated harmonisation toolboxes like FedHarmony ^45^, which should be considered in future research for optimal model performance.

### The road ahead for real-world FL in neuroimaging

Federated learning offers a compelling alternative to centralised machine learning. It distributes computational load and addresses data sharing as one of the major impediments for international scientific collaborations (47). The promises for federated learning in the medical field are big indeed, and especially come from cancer research. One of the first large-scale real-world FL demonstrations was in 2022 by Sarthak Pati and colleagues (15). In a global federated learning setting with 71 institutions, they updated a publicly available algorithm to detect glioblastoma boundaries and obtained a considerable gain in the validation Dice score across clients of about 25%. The authors mention “It is the use of FL that successfully enabled (i) access to such an unprecedented dataset of the most common and fatal adult brain tumour, and (ii) meaningful ML training to ensure the generalizability of models across out-of-sample data.”. Indeed, although Vo et al. 2024 confirms that comparable results can be obtained in centralised contexts in ideal scenarios (48), the key benefit of FL is that it can still achieve these results when this is not the case. Similarly, technical advances enable addressing data heterogeneity across institutions in a federated setting, such as tackling the non-IID issue by a combination of distributed gradient blending and proximity-aware client weighting (49). Considering these medical FL breakthroughs, it is even more surprising that literature appears to ignore the very first step to achieve these results: how to build an FL network across clinical institutions. This practical step appears to be an assumption, as literature focusses on challenges after a network has been established (50). In our experience however, building a real-world FL network is the key roadblock between the compelling idea of federated learning and its practical realisation, and should be the key focus to start an era of real-world decentralised machine learning beyond simulation.

#### Collaboration

A first prerequisite is a minimal trust between all partners in the FL network. With malicious intent, source data could for example be shared between computers via VPN. The team should moreover be multidisciplinary, containing both medical experts and technical experts in terms of information technology (IT) and data science. Real-world federated learning poses unique IT challenges in terms of communication and encryption on top of other existing technical challenges in deep learning research. On the other hand, medical expertise is required in preprocessing and interpreting the data, as well as understanding the model and its output.

#### Hardware

Besides sufficient storage, updating neural networks like the DenseNet model (over 11 million parameters) requires processing units like a graphical (GPU) or tensor processing unit (TPU) on each client computer. In our network, all client computers were equipped with a GPU of 24GB random access memory (RAM).

#### Software

There is a lack of FL toolboxes that are rigorously tested in real-world circumstances, and most are designed as general frameworks not specifically tailored to medical data needs. We started designing FLightcase in early 2023 when real-world FL examples in neuroimaging just started to emerge (15). Although some toolboxes such as Flower (13) and OpenFL (51) were distributed open source, we experienced difficulties in real-world deployment on our own network. We therefore set out to design a framework relying on a simple but stable connection between computers, focussing on the core needs of our methodology, i.e., compliance with BIDS, supporting loading anatomical brain images and focussing on the original and most popular FL algorithm FedAvg. In the meantime, toolboxes such as Flower have further developed into communities with peer support. Awareness on the difficulty of real-world FL is rising, reflected by companies facilitating deployment such as ScaleOut, and forces are joined to allow tailoring toolboxes to the specific needs of different modelling domains. Almost a decade after the pioneering work of Sergey M. Plis and colleagues (52) on standardised analysis of decentralised neuroimaging data, FL software approaches a technology readiness level where centralised workflows can seamlessly be converted to federated ones. For FLightcase specifically, we in the meantime reconsidered the basic communication layer, allowing file sharing by uploading to and downloading from a Flask web server.

#### Connectivity and remote access

All computers in our network are connected to locally available internet providers. This introduces new centre-specific difficulties pertaining to local security settings. Blocking web addresses such as Teamviewer for example complicated remote access in our case. Combined with limited scalability due to a maximum number of remote computers in TeamViewer, we switched to tmate (cfr. methods). The drawback of this approach is that there is no graphical interface available; a remote computer is operated via command line.

#### Financial

Ensuring the aforementioned requirements comes with a price tag. Especially the hardware for each client with expensive GPUs imposes additional financial restraints, limiting the feasibility of setting up a network.

## Data and code availability

The source code is available in the GitHub repository of the AIMS lab (https://github.com/AIMS-VUB/FLightcase)(53), whereas FLightcase was also distributed via the Python Package Index (PyPI). Data underlying the real-world and simulated brain age experiments is open source available.

## Author contributions

Conceptualization: SD, JL, MDV, OYC, JVS, DMS, GN; Data curation: SD; Formal Analysis: SD; Funding acquisition: SD, MV, DH, JB, JVS, DMS, GN; Investigation: SD, MG, MV, TU, JL, MKu, DH, JB, IKP, MKi, JM, JVS, DMS, GN; Methodology: SD, JL, MKu, JB, OYC, JVS, DMS, GN; Project administration: TU, MV, DH, OYC, JVS, DMS, GN; Resources: MG, MV, TU, MKu, DH, IKP, MKi, JM, JVS, GN; Software: SD, JL; Supervision: JVS, DMS, GN; Validation: SD; Visualization: SD; Writing – original draft: SD; Writing – review & editing; all but SD.

## Data Availability

https://cam-can.mrc-cbu.cam.ac.uk/dataset/

https://brain-development.org/ixi-dataset/

https://fcon_1000.projects.nitrc.org/indi/retro/sald.html

## Acknowledgements

This study was funded by a personal industrial PhD grant (Baekeland, HBC.2019.2579) appointed by Flanders Innovation and Entrepreneurship to Stijn Denissen, a personal travel grant (V412023N) appointed by the “Fonds Wetenschappelijk Onderzoek” (FWO) to Stijn Denissen for his stay in Prague in the context of this study, a grant (SRP85) from the Vrije Universiteit Brussel and a junior post-doctoral grant (12A6U25N) from the FWO Flanders. This project was furthermore funded by an IOF-POC grant (IOFPOC57) from the Vrije Universiteit Brussel (VUB), by an ITEA grant (20030 HeKDisco, HBC.2021.0500) from Flanders Innovation and Entrepreneurship, by an institutional support from the Czech Ministry of Health (RVO-VFN 64165) and by the National Institute for Neurological Research, Czech Republic, Programme EXCELES, ID Project No. LX22NPO5107, funded by the European Union – Next Generation EU. Guy Nagels is a senior clinical research fellow of the FWO Flanders (1805620N).

We would like to thank Jelle Laton and Robert Malinowski for their help in the setup and maintenance of the hardware present in the federated learning network. We also thank André Vital Serafim Silva for the insights in centralised brain age modelling generated in his master thesis. The following tools were used for figure creation: the Mac desktop version of draw.io (https://www.draw.io/), adapted icons from JGraph (https://github.com/jgraph, licensed under CC BY 4.0 (https://creativecommons.org/licenses/by/4.0/)), flag icons from flagcolorcodes (https://www.flagcolorcodes.com/) and the Python package GADM v0.0.3 (54).

