## supplementary material for "Real-world federated learning for the brain imaging scientist"

#### FLightcase unpacked

FLightcase has a command line interface (CLI) with three main functionalities. All functionalities require the definition of one argument, which is the path to the FL workspace.

- **prepare-workspace** This will start a procedure to be completed in the command line, creating the FL workspace with the necessary files. This is only a settings JSON for the client, while an FL plan JSON and model architecture Python file are also prepared for the server workspace. Details stored in the JSON files are described in supplementary table S1 (server settings), S2 (client settings) and S3 (FL plan). The first option is to only copy the template files to the workspace folder, and to complete it separately with a text editor. The second option is to fill in the template file in the command line. Only JSON files can be filled in this way, the template architecture Python file is automatically copied to the server workspace (cfr. option 1).
- **run-server** This will run the server.py Python script, defining all steps to be carried out by the server. Its steps are described visually in the left panel of figure S1.
- **run-client** This will run the client.py Python script, defining all steps to be carried out by a client. Each client in the federation runs this command to participate. Its steps are described visually in the right panel of figure S1.

Below, the stepwise interplay between the server.py and client.py script is explained narratively, while we refer to figure S1 for the visual overview. The code is based on the Federated Averaging (FedAvg) algorithm (1).

**Step 1: The server starts the FL experiment.** After having prepared the FL workspace at each node, and all computers joined the federation by either running run-server or run-client, the FL process starts. As can be verified in figure S1, the order does not matter.

Assuming the server started first, it will first extract the settings (cfr. supplementary table S1) that define [1] the server workspace path, [2] which clients the server can expect to join the federation and [3] whether the server needs to use an existing set of weights, a "state", as initial model (default torch initialisation is used otherwise). The server will then wait for the client dataset sizes to arrive.

**Step 2: Clients join the FL experiment.** When a client joins the federation, it will first extract its settings JSON (cfr. supplementary table S2). It will then load the “participants.tsv” file containing the label to predict and append columns containing the paths to the brain information as input for the model. The client then sends its dataset size to the server and waits for the FL plan and the model architecture. After extraction of the FL plan, the client prepares for deep learning (e.g. defining the optimizer) and waits for the global model to be sent by the server.

**Step 3: The server prepares the global model.** When all clients have joined the federation and informed the server on their dataset sizes, the server sends the FL plan and model architecture to all clients, and extract the FL plan for the FL orchestration. It then prepares the initial global model, which is either taken from an existing set of weights (cfr. supra) or initialised from scratch. The meta-info of the model (e.g. number of parameters) is saved to the server workspace, and after sending the initial model to all clients, it waits for the updated, "local" client models.

**Step 4: The client updates the global model.** Upon receiving the initial global model, the client starts training it on the local dataset. It creates a subfolder to store model states in, loads the global model and trains/validates its model given a predefined number of bootstraps defined in the FL plan. The training results (e.g. loss) of all bootstraps is sent to the server, and a weighted average across all bootstrap models (weight is the reciprocal of the loss (1/loss)) yields a local model that is sent to the server. If the average loss across the bootstraps is smaller than the minimum value, this round was successful and there is no need to update the learning rate. Its counter is therefore reset to 0, and a new minimum average loss is stored. If it is higher than or equal to the current average loss, the counter increases with one (training stagnates). If the predefined (FL plan) number of rounds without improvements is met, the learning rate is reduced. The client then waits again for the new global model to arrive.

**Step 5: The server aggregates the client models.** Upon reception of all local client models, the server aggregates the model by performing a weighted average of a random subset of local models; models that were trained on larger data sets receive a higher weight. Based on the average validation loss of the clients in the subset for this federated learning round, the model either labels the model as the best model yet or increases a counter of the number of FL rounds without improvement. When a predefined number is met, this will send a text file that prompts all clients to exit the training phase, and wait for the final model. If this is not the case, the new global model is sent to all clients, who perform step 4 again.

**Step 6: Sending and testing the final model.** If the predefined number of FL rounds have been performed or the FL process was aborted prematurely by the server, the server sends the final, best model (lowest average validation loss) to all clients, who are waiting for it. The clients then apply this model to their test data and share their results with the server. The server in the meantime awaits this information and saves it. The program is then stopped by all nodes in the FL network.

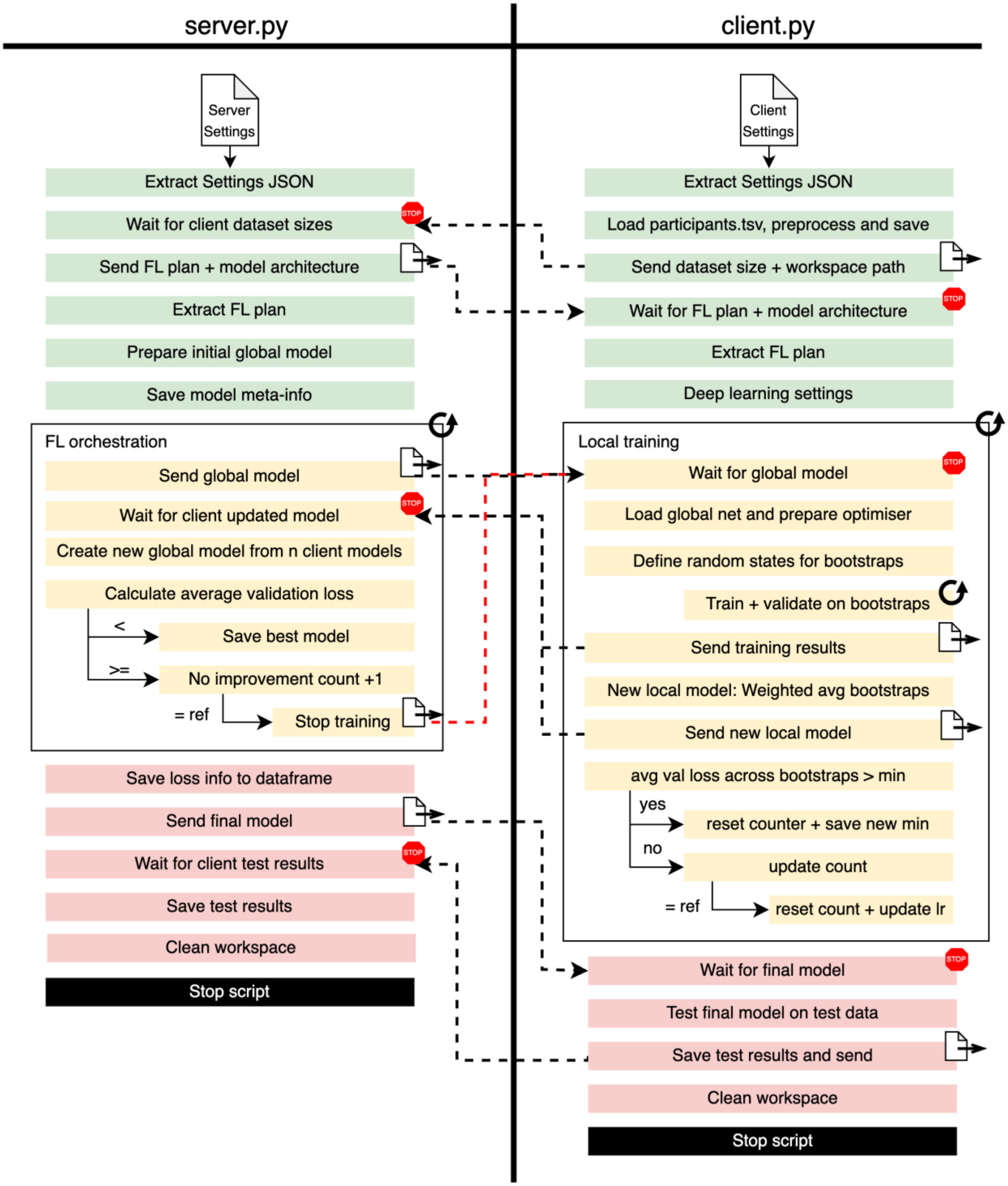


*Figure S1. Visual description of the interaction between the server.py and client.py scripts.*

### How to test FLightcase

This section is dedicated to testing FLightcase in a lightweight environment, allowing any scientist run the simulation without high computational and storage resources. The simulation is a smaller brain age experiment that can be found in the “simulation” subfolder of the FLightcase GitHub repository (https://github.com/AIMS-VUB/FLightcase). The simulation was tested on a Mac laptop:

- **Hardware.** Apple MacBook Pro M3, 11 CPU cores, 14 GPU cores, 18GB random access memory (RAM) and a 16-core Neural Engine (designed to accelerate AI and ML tasks).
- **Operating system.** Sonoma v14.1
- **Python version:** 3.12.7
- **Git version:** 2.39.3

**Data.** The simulation relies on three OpenNeuro BIDS datasets (2–4) from three studies on healthy participants (5–7). The datasets contain age-labelled T1-weighted brain MR images. Data preparation is performed in a conda environment (8), which is created with the "environment.yml" file (python version: 3.11). It first consists of downloading the data via the CLI of Amazon Web Services (AWS). The data is downloaded in an "inputs" subfolder of the FLightcase GitHub repository, which is ignored by Git. After pulling the contents of the submodule with "git submodule init" and "git submodule update", the T1-weighted MR images are pre-processed using the pipeline by Wood et al. 2022. Finally, the images are downsampled to 26 voxels isotropically to reduce computational load for the simulation.

**Network architecture.** A lightweight CNN was defined to maximally reduce the computational load of the local simulation, which is an adapted version of the Simple Fully Convolutional Network (SFCN) (9). The feature extractor consists of a succession of three convolutional blocks, each consisting of a convolutional layer, a 3D batch normalisation, a max pooling layer (only the first 2 blocks) and a rectified linear unit (ReLU). The number of channels inflates from 1 to 32 to 64 to 128. The classifier reduces the 128 feature maps to a single scalar value, and consists of an average pool, dropout, flatten and linear layer. The total network has only 65153 parameters.

**Running the simulation.** All practical steps are described in the “README.md” file in the “simulation” subfolder. As the simulation includes the server and three clients, four workspace directories are to be prepared using the "FLightcase prepare-workspace" command (cfr. supra). For all settings JSON files, "127.0.0.1" (localhost) can be used as IP address. When this IP address is used, the “cp” command is used to communicate instead of the “scp” command, as a persistent exception was raised in a simulated environment by “paramiko” (10). We recommend installing FLightcase in a virtual or conda environment, after which it can be activated in four different terminals: 1 for the server and 3 for the clients. Using the "run-server" and "run-client" commands with the path to the settings JSON, the FL process starts.

| Key | Value |
|---|---|
| "workspace_path_server" | "/path/to/server/workspace/repository" |
| "initial_state_dict_path" | null |
| "client_credentials" | {<br> “name_client_1”: {<br> "ip_address": "ip_address_client_1",<br> "username": "username_client_1",<br> "password": "password_client_1"<br> },<br> “name_client_2”: {<br> "ip_address": "ip_address_client_2",<br> "username": "username_client_2",<br> "password": "password_client_2"<br> }<br>} |

*Table S1. Template of server settings JSON file.*

| Key | Value |
|---|---|
| "workspace_path_client" | "/path/to/client/workspace/repository" |
| "client_name" | "name_of_client_computer" |
| "server_ip_address" | "127.0.0.1" |
| "server_username" | "username_server_computer" |
| "server_password" | "password_server_computer" |
| "workspace_path_server" | "/path/to/server/workspace/repository" |
| "derivative_name" | null |
| "modalities_to_include" | {<br> "anat": ["T1w.nii.gz", "FLAIR.nii.gz"]<br>} |
| "colnames_dict" | {<br> "id": "subject_id_BIDS",<br> "session": "session_BIDS",<br> "label": "label"<br>} |
| "subject_sessions" | null |
| "bids_root_path" | "/path/to/BIDS/root/directory/" |
| "batch_size" | 8 |

*Table S2. Template of client settings JSON file.*

| Key | Value | Explanation |
|---|---|---|
| "n_rounds" | 5 | Number of federated learning rounds |

| | | |
|---|---|---|
| "n_epochs_per_round" | 1 | One epoch is one complete model update on all available training data. |
| "lr" | 0.001 | The learning rate controls how the model's weights are updated based on the gradient, namely by controlling the magnitude of the step taken into the opposite direction of the gradient. |
| "pat_lr_red" | 3 | The number of FL rounds without validation loss improvement (tracked per client) before reducing the learning rate by the learning rate reduction factor (cfr. infra). |
| "pat_stop" | 10 | Number of FL rounds without improvement of the average validation loss across clients before stopping training early. |
| "lr_reduce_factor" | 0.5 | Factor by which the learning rate is reduced after several rounds (cfr. supra) without validation loss improvement (tracked per client). |
| "train_fraction" | 0.6 | Fraction of client data used for training |
| "val_fraction" | 0.2 | Fraction of client data used for validation |
| "test_fraction" | 0.2 | Fraction of client data used for testing |
| "n_clients_set" | 2 | Number of clients of which the local model is used for aggregation to a new global model |
| "n_splits" | 1 | Number of random train/validation splits (using bootstrapping) for each FL round. |
| "criterion" | "l1loss" | Loss function. In this case, this is the L1 loss, $\sum_{i=1}^{n}\lvert y_i - \hat{y}_i\rvert$. |
| "optimizer" | "adam" | Optimiser algorithm for updating model parameters. We used Adam optimisation (11). |

*Table S3. Template federated learning plan*

| Key | Value |
|---|---|
| "n_rounds" | 300 |
| "n_epochs_per_round" | 1 |
| "lr" | 0.0001 |
| "pat_lr_red" | 3 |
| "pat_stop" | 20 |
| "lr_reduce_factor" | 0.5 |
| "train_fraction" | 0.6 |
| "val_fraction" | 0.2 |
| "test_fraction" | 0.2 |
| "n_clients_set" | 2 |
| "n_splits" | 5 |
| "criterion" | "l1loss" |

| | |
|---|---|
| "optimizer" | "adam" |

*Table S4. Federated learning plan for the real-world experiments*

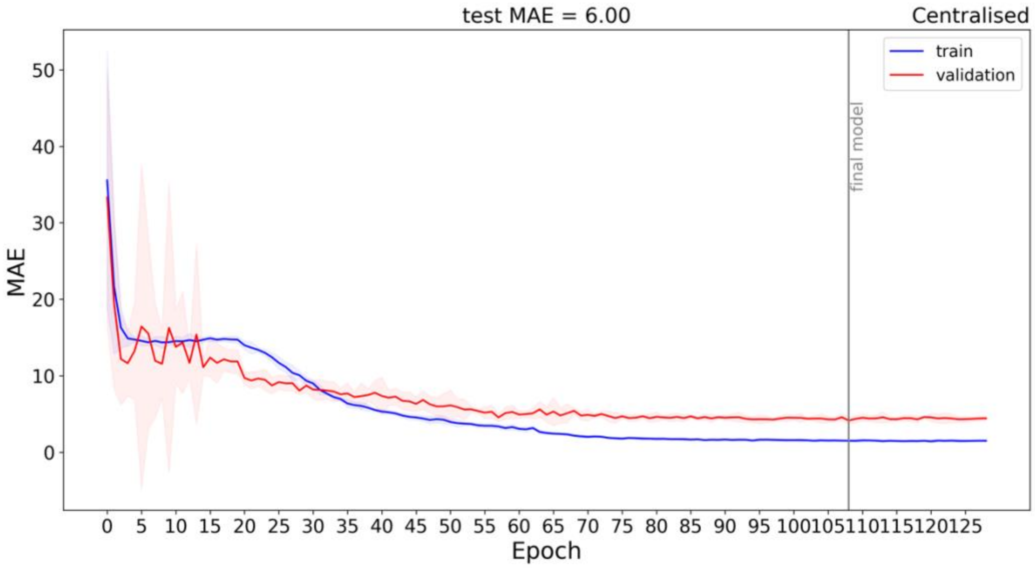


*Figure S2: Centralised training of the brain age network. The red (train) and blue (validation) lines represent the average train and validation MAE respectively, and the shaded red and blue areas represent the 95% confidence interval, all calculated across 5 bootstraps per epoch.*

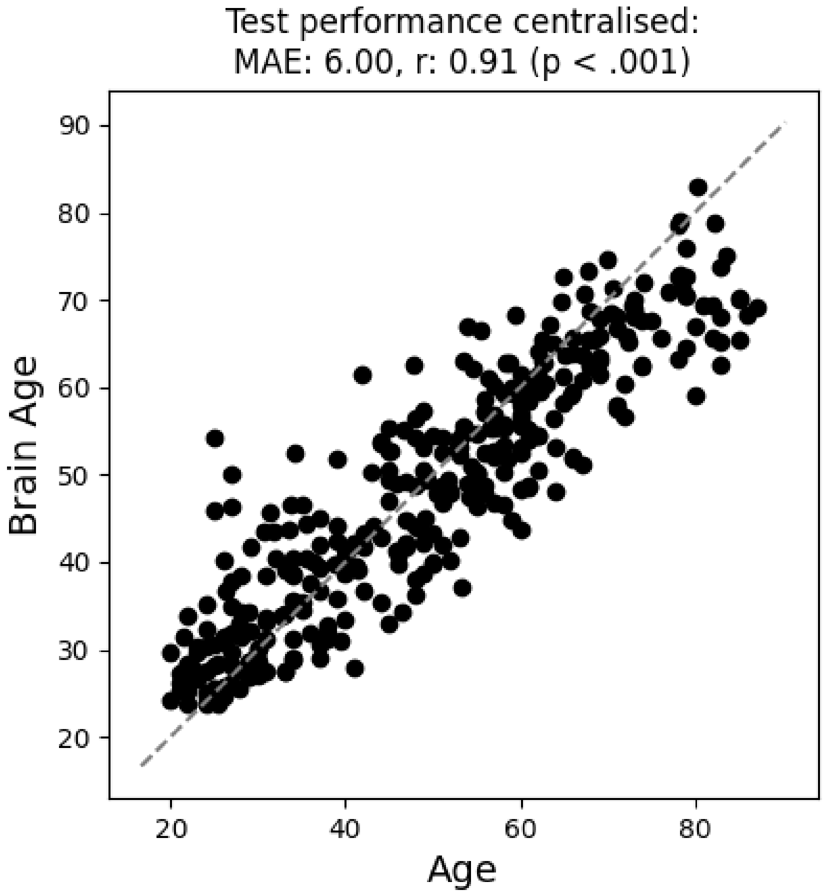


*Figure S3. Centralised brain age test results. The grey dashed line indicates the perfect prediction line where Age = Brain Age.*
